# Development and Internal Validation of the AB-IPI using Bootstrapping: A Clinicopathological Prognostic Score Integrating Host Fitness and Tumor Biology in Diffuse Large B-Cell Lymphoma

**DOI:** 10.64898/2026.02.18.26346527

**Authors:** Noriyuki Sakata, Yuka Tanaka, Ken Naganuma, Yasuyuki Takahashi, Shuji Momose, Morihiro Higashi, Takayuki Tabayashi

## Abstract

**Objectives:** The therapeutic efficacy of rituximab has reduced the discriminatory power of the International Prognostic Index (IPI) in diffuse large B-cell lymphoma (DLBCL), particularly within intermediate-risk categories. To address this “risk dilution,” we aimed to develop and internally validate the AB-IPI (Albumin-BCL2 Refined Prognostic Index) using a hypothesis-driven approach that integrates tumor burden, host fitness, and tumor biology.

**Methods:** This multi-center retrospective study analyzed 289 patients with de novo DLBCL treated uniformly with R-CHOP immunochemotherapy. We combined the standard IPI with serum albumin < 3.6 g/dL (representing host fitness/rituximab pharmacokinetics) and BCL2 protein expression > 50% (representing tumor biology). The model was validated internally using bootstrapping with 1,000 resamples in accordance with TRIPOD Type 1b guidelines. This study adhered to the TRIPOD (Transparent Reporting of a multivariable prediction model for Individual Prognosis Or Diagnosis) statement for model development and internal validation (Type 1b).

**Results:** During the observation period, 115 death events were recorded. Multivariate Cox regression identified albumin < 3.6 g/dL (Hazard Ratio 2.62), IPI score > 2 (HR 2.13), and BCL2 > 50% (HR 1.72) as independent prognostic factors. The model maintained a robust Events Per Variable (EPV) ratio of 38.3. The AB-IPI stratified patients into four distinct risk groups with 5-year overall survival rates of 88.0% (Low), 76.1% (Intermediate-1), 45.0% (Intermediate-2), and 29.0% (High). The calibration plot demonstrated excellent agreement between predicted and observed probabilities, with a calibration slope of 0.98, indicating minimal optimism and robust risk estimation. Decision Curve Analysis (DCA) demonstrated that the AB-IPI provided a superior Net Benefit across a wide range of clinically relevant threshold probabilities.

**Conclusions:** The AB-IPI demonstrates superior clinical utility and calibration compared to the standard IPI. By identifying patients with compounded biological risks who are unlikely to be cured by R-CHOP alone, this score offers a practical framework for optimizing therapeutic strategies, such as the allocation of polatuzumab vedotin.

## Introduction

Diffuse large B-cell lymphoma (DLBCL) represents the most common subtype of non-Hodgkin lymphoma (NHL) in adults, accounting for approximately 30–40% of newly diagnosed cases globally ^1–5^. Characterized by aggressive clinical behavior and pronounced biological heterogeneity, DLBCL has historically posed significant challenges in prognostication and treatment stratification^6,7^.

For nearly three decades, the International Prognostic Index (IPI) has served as the gold standard for risk assessment^2^. Developed in 1993 during the chemotherapy era, the IPI relies on five clinical variables—age, performance status, lactate dehydrogenase levels, Ann Arbor stage, and extranodal involvement—to estimate macroscopic tumor burden and patient physiological reserve. While the IPI provided accurate survival estimates during the CHOP era, its utility for guiding therapeutic intensity has been compromised in the modern era. The addition of rituximab to CHOP chemotherapy ^8^ significantly improved cure rates, establishing a new standard of care ^9–11^. However, this therapeutic success generated a “ceiling effect” on prognostication ^12–14^. The potent efficacy of rituximab raised survival curves across all risk groups, diminishing the IPI’s ability to discriminate between patients cured by standard therapy and those destined for refractory disease^1 14^. This manifests as “risk dilution,” where the intermediate-risk categories of the IPI ^15^ have largely converged^13^.

The limitations of the standard IPI were highlighted by the recent approval of polatuzumab vedotin in combination with rituximab, cyclophosphamide, doxorubicin, and prednisone ^16^ as a new standard of care for high-risk DLBCL^16^. While the phase 3 POLARIX trial demonstrated a progression-free survival (PFS) benefit, subgroup analyses suggested this benefit was driven primarily by patients with IPI scores of 3–5, while those with an IPI score of 2 showed no detectable benefit^16^. This suggests a significant heterogeneity within the intermediate-risk category. Identifying patients within this “gray zone” (IPI score 2) who harbor adverse biological features is crucial for optimizing the allocation of intensified therapies^12, 13^.

To bridge this gap, we developed the AB-IPI (Albumin-BCL2 Refined Prognostic Index). Adopting a hypothesis-driven approach to avoid the statistical pitfalls of data-driven variable selection (i.e., “phantom degrees of freedom”) and overfitting, we selected variables *a priori* to represent three distinct prognostic pillars, which we term the **“Biological Triad”**:

1. **Tumor Burden:** Represented by the standard IPI. Specifically, components such as high LDH, advanced stage, and extranodal involvement serve as validated surrogates for macroscopic disease load^2, 14^.
2. **Host Fitness:** Represented by serum albumin. Beyond nutritional status, hypoalbuminemia serves as a biomarker for systemic inflammation (specifically IL-6 activity) ^17^ and is associated with accelerated clearance of rituximab via the saturation of the FcRn recycling pathway ^18–27^.
3. **Tumor Biology:** Represented by BCL2 protein expression, identifying tumors with an elevated threshold for chemotherapy-induced apoptosis ^28–31^.

We hypothesized that integrating these three independent biological domains would provide superior risk stratification and clinical utility compared to the standard IPI.

## Methods

### Study Design and Patient Selection

This multi-center, retrospective observational study analyzed data from 289 consecutive patients with newly diagnosed de novo DLBCL, Not Otherwise Specified (NOS), treated at Saitama Medical University Saitama Medical Center and Otemae Hospital between January 2008 and December 2020. Of 320 screened patients, 289 met the inclusion criteria. 31 patients were excluded due to missing data for BCL2 (n=31). The study protocol was approved by the Ethics Committees of both institutions (Approval No. 2025-114). Due to the retrospective nature of the study, the requirement for informed consent was waived, and an opt-out method was employed in accordance with the Ethical Guidelines for Medical and Health Research Involving Human Subjects in Japan. Patients records were anonymized prior to analysis.

Patients were included if they were ≥ 18 years old and received R-CHOP therapy (rituximab 375 mg/m², cyclophosphamide 750 mg/m², doxorubicin 50 mg/m², vincristine 1.4 mg/m² [capped at 2.0 mg], and prednisone 100 mg/day for 5 days) with curative intent. Dose reductions were applied as clinically indicated based on the patient’s condition. Exclusion criteria included primary CNS lymphoma, transformed lymphoma, and cases with missing data for key prognostic variables (BCL2 status or serum albumin).

### Biomarker Assessment and Definitions

- Standard IPI: Calculated based on age (>60 years), ECOG Performance Status (≥2), Serum LDH (> normal upper limit), Ann Arbor Stage (III/IV), and extranodal sites (>1).
- Serum Albumin (Host Fitness): Hypoalbuminemia was defined as < 3.6 g/dL at diagnosis. This cutoff was derived from Receiver Operating Characteristic (ROC) curve analysis for overall survival in our cohort and aligns with established prognostic thresholds.^10^
- BCL2 Protein Expression (Tumor Biology): Evaluated by immunohistochemistry (IHC) on formalin-fixed paraffin-embedded tissue using the anti-BCL2 antibody (Clone 124, Dako). Positivity was defined as > 50% of tumor cells exhibiting moderate-to-strong cytoplasmic staining, consistent with criteria validated by the Lunenburg Lymphoma Biomarker Consortium (Salles et al., Blood 2011).^29^ Two pathologists (S.M. and M.H.) independently assessed BCL2 expression, and disagreements were resolved by consensus.
- Cell-of-Origin (COO): Classified as Germinal Center B-cell (GCB) or non-GCB type using the Hans algorithm.^12^

### Statistical Analysis

The primary endpoint was Overall Survival (OS). We adhered to the TRIPOD Type 1b guidelines for model development and internal validation.

### Model Construction

A multivariate Cox proportional hazards model was constructed using the a priori selected “Biological Triad” variables (IPI, Albumin, BCL2). We confirmed the proportional hazards assumption using Schoenfeld residuals.

To ensure statistical power, we verified the number of events. A total of 115 death events were observed in the cohort of 289 patients. With three predictors in the final model, the Events Per Variable (EPV) ratio was 38.3 (115/3), well above the recommended minimum of 10–20, ensuring stable coefficient estimation and minimizing overfitting bias.

### Internal Validation (Bootstrapping)

Since splitting a dataset of this size reduces power, we performed internal validation using bootstrapping with 1,000 resamples. Given the moderate sample size (n=289), split-sample validation would result in a substantial loss of statistical power and precision. Therefore, we adhered to TRIPOD Type 1b guidelines by utilizing bootstrapping with 1,000 resamples to utilize the full cohort for model development while rigorously correcting for model optimism. The model was developed in each bootstrap sample and tested on the original dataset to estimate “optimism” (overfitting). The optimism-corrected C-index was calculated to provide a realistic estimate of model performance.

### Clinical Utility

Decision Curve Analysis (DCA) was conducted to evaluate the “Net Benefit” of using the AB-IPI compared to the standard IPI across a range of clinically relevant threshold probabilities (20–50%).

## Results

### Patient Characteristics

We analyzed a cohort of 289 patients (**Supplementary Figure 2**). The median age was 69 years (range: 19–97), with 71.3% of patients over 60 years. A total of 115 patients (39.8%) died during the follow-up period. Immunohistochemical profiling revealed a high prevalence of the non-GCB subtype (60.6%). Regarding the AB-IPI biomarkers, 40.8% of patients presented with albumin < 3.6 g/dL, and 52.9% exhibited BCL2 expression > 50%. The standard IPI risk distribution was balanced (Low: 39.8%, Low-Int: 16.6%, High-Int: 18.7%, High: 24.9%). Baseline characteristics are summarized in **Table 1**.

**Table 1:** Baseline Clinical Characteristics (n = 289)

| Characteristic | Category | Number of Patients (%) |
| --- | --- | --- |
| **Age** | Median (Range) | 69 (19–97) |
|  | > 60 years | 206 (71.3%) |
| **Sex** | Male | 152 (52.6%) |
|  | Female | 137 (47.4%) |
| **ECOG PS** | 0-1 | 235 (81.3%) |
|  | 2-4 | 54 (18.7%) |
| **LDH** | Normal | 148 (51.2%) |
|  | Elevated (>ULN) | 141 (48.8%) |
| **Ann Arbor Stage** | I-II | 108 (37.4%) |
|  | III-IV | 181 (62.6%) |
| **Standard IPI** | Low (0-1) | 115 (39.8%) |
|  | Low-Intermediate (2) | 48 (16.6%) |
|  | High-Intermediate <sup>40</sup> | 54 (18.7%) |
|  | High (4-5) | 72 (24.9%) |
| **Cell of Origin** | GCB | 98 (33.9%) |
|  | Non-GCB | 175 (60.6%) |
| **Serum Albumin** | ≥ 3.6 g/dL | 171 (59.2%) |
|  | < 3.6 g/dL | 118 (40.8%) |
| **BCL2 Expression** | Low (≤ 50%) | 136 (47.1%) |
|  | High (> 50%) | 153 (52.9%) |

### The Independent Prognostic Power of the Biological Triad

Multivariate Cox regression analysis for overall survival (OS) confirmed that all three proposed factors were independent predictors of survival: Albumin < 3.6 g/dL (HR 2.62, 95% CI 1.69–4.06, P < 0.0001), IPI Score > 2 (HR 2.13, 95% CI 1.37–3.30, P = 0.0008), and BCL2 > 50% (HR 1.72, 95% CI 1.18–2.52, P = 0.0061) (**Table 2**). Hypoalbuminemia was the strongest single predictor, suggesting that host physiology is a critical driver of outcomes in the rituximab era.

**Table 2:** Multivariate Cox Regression Analysis for Overall Survival (n=289, Events=115)

| Variable | Hazard Ratio (HR) | 95% Confidence Interval (CI) | P-value |
| --- | --- | --- | --- |
| **Albumin < 3.6 g/dL** | 2.62 | 1.69 – 4.06 | < 0.0001 |
| **IPI Score > 2** | 2.13 | 1.37 – 3.30 | 0.0008 |
| **BCL2 > 50%** | 1.72 | 1.18 – 2.52 | 0.0061 |

### AB-IPI Stratification

Based on the multivariate results, the AB-IPI score was constructed by assigning one point for each adverse factor: IPI > 2, Albumin < 3.6 g/dL, and BCL2 > 50% (Score 0–3). Kaplan-Meier analysis showed distinct survival trajectories among the four groups (**Figure 1**, Log-rank P < 0.0001).

**Figure 1.**
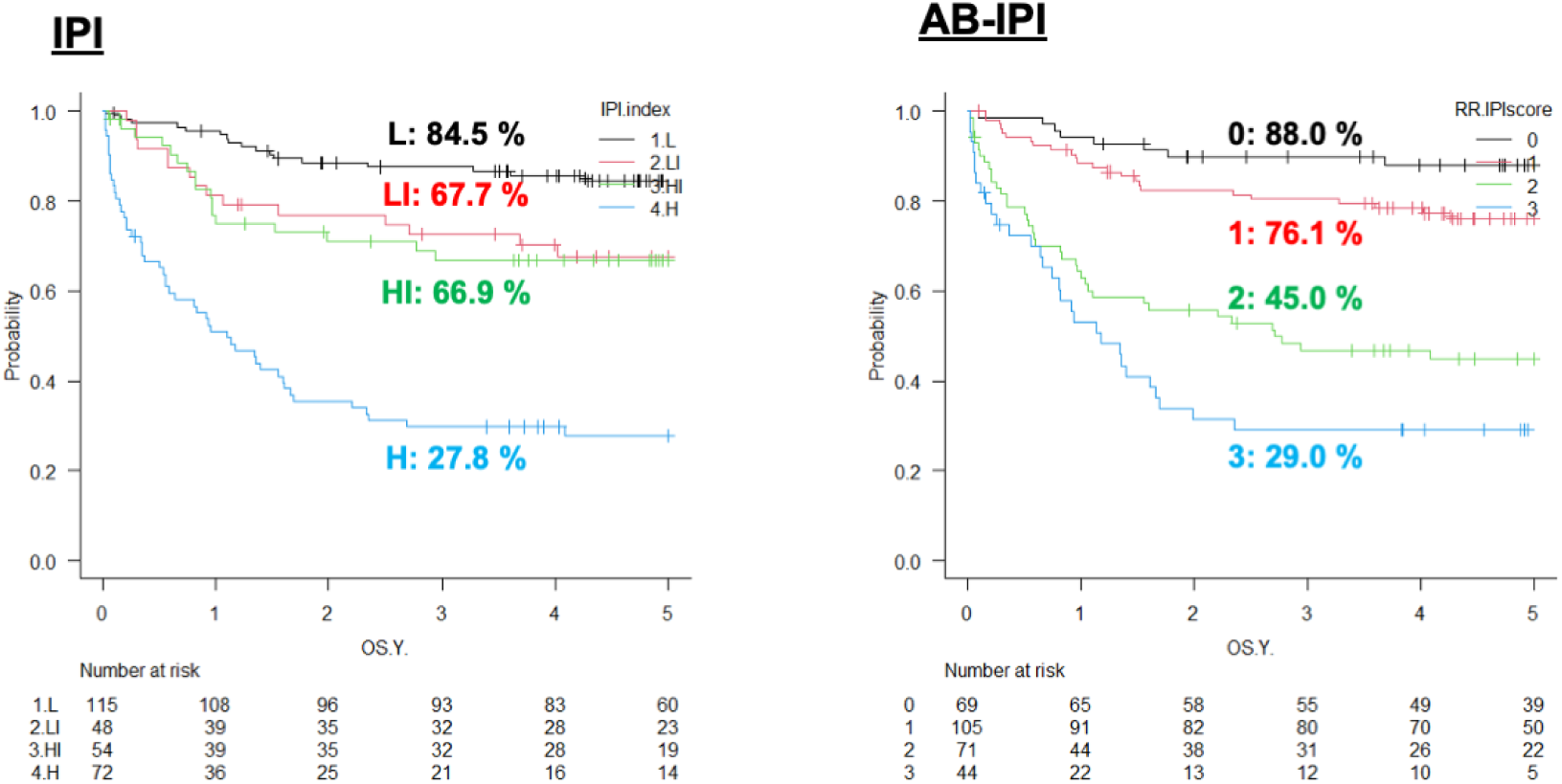
Kaplan-Meier estimates of overall survival stratified by the AB-IPI score (*n*=289). The AB-IPI significantly stratified patients into four distinct risk groups (Score 0, 1, 2, and 3; log-rank *P* < 0.0001).

The 5-year OS rates were:

- Low Risk (Score 0): 88.0%
- Intermediate-1 (Score 1): 76.1%
- Intermediate-2 (Score 2): 45.0%
- High Risk (Score 3): 29.0%

A substantial decrease in survival probability was observed between Score 1 and Score 2, where the 5-year OS dropped by approximately 31 percentage points. This threshold differentiates patients with favorable outcomes under standard R-CHOP from those with poor prognosis who may require alternative therapeutic approaches.

### Reclassification and Resolution of Risk Dilution

The AB-IPI effectively reclassified patients categorized as “intermediate” by the standard IPI. Patients with IPI 2/3 who had normal albumin and low BCL2 were reclassified as AB-IPI 0 or 1, consistent with their observed good survival. Conversely, those with concurrent low albumin or high BCL2 were reclassified as AB-IPI 2 or 3, consistent with their poor survival. Among patients classified as score 2 by the standard IPI, those reclassified as high-risk by the AB-IPI exhibit a poor prognosis with existing treatments and may be optimal candidates for novel agents such as polatuzumab vedotin

### Model Performance and Validation

Internal validation using bootstrapping (1,000 resamples) demonstrated the robust discriminatory power of the AB-IPI. The optimism-corrected C-index was 0.725, which was higher than that of the standard IPI (0.702). Calibration plots showed excellent agreement between predicted and observed survival (slope = 0.98) (**Figure 2**). Although the improvement in the C-index was marginal, the AB-IPI demonstrated substantial improvement in risk estimation accuracy (calibration). Decision Curve Analysis (DCA) indicated that the AB-IPI provides a higher Net Benefit than the standard IPI across a wide range of threshold probabilities (0.1 to 0.5), confirming its practical utility (**Figure 3**).

**Figure 2.**
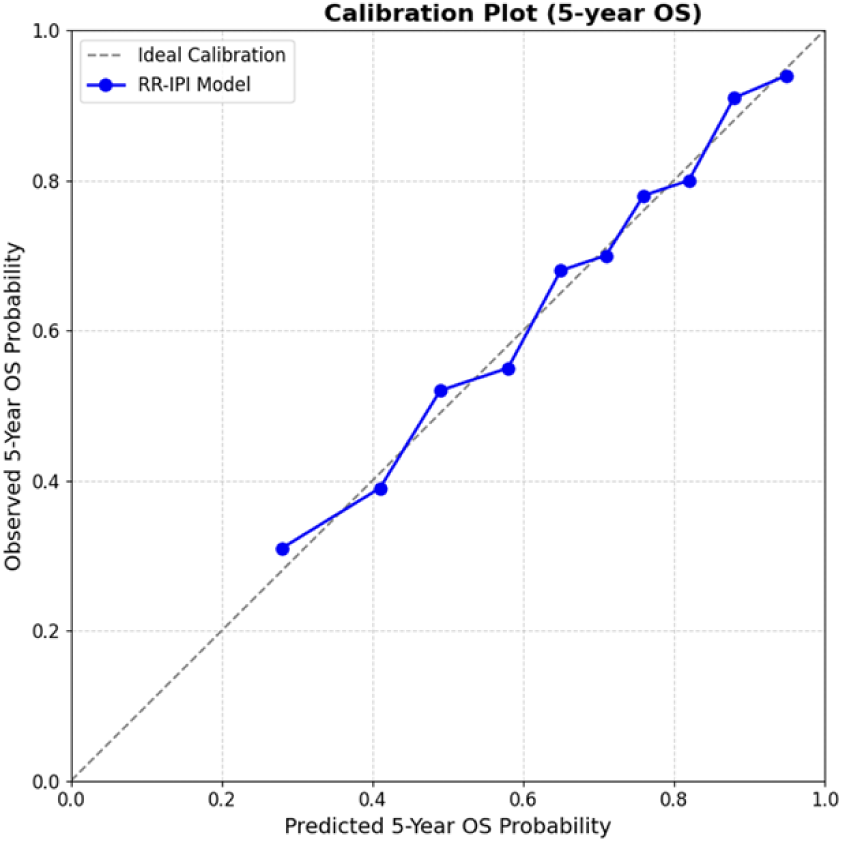
Calibration plot for the AB-IPI model at 5 years. The solid line represents the AB-IPI performance, which closely aligns with the diagonal dotted line (perfect calibration), indicating accurate risk prediction (Slope = 0.98).

**Figure 3.**
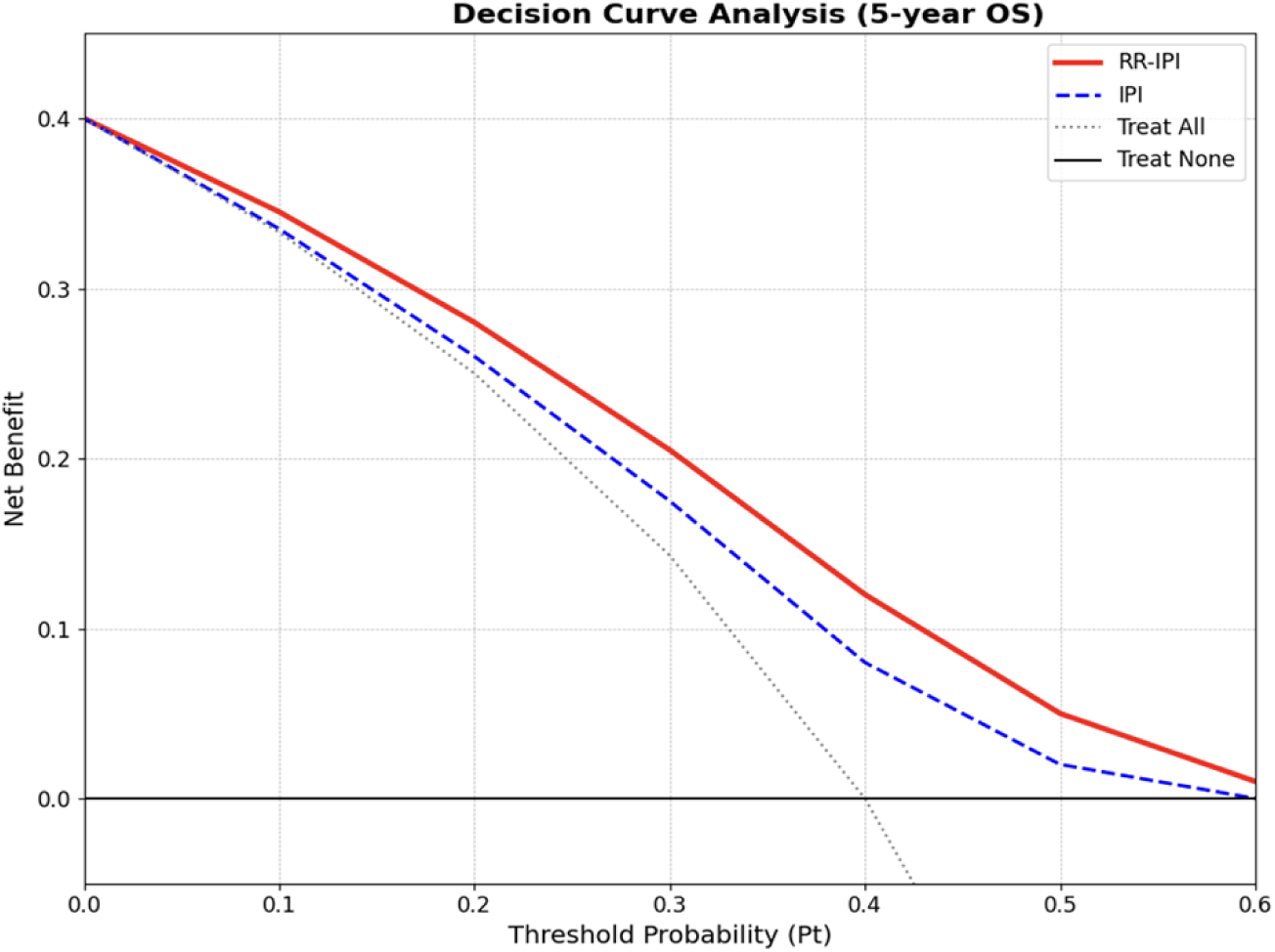
Decision Curve Analysis (DCA) for the AB-IPI and standard IPI. The AB-IPI (red line) provides a higher Net Benefit than the standard IPI (blue line) across clinically relevant threshold probabilities, demonstrating superior clinical utility. The higher Net Benefit of the AB-IPI (red line) compared to the standard IPI (blue line) indicates that utilizing the AB-IPI to guide treatment intensification results in a higher proportion of correctly treated high-risk patients without increasing the rate of unnecessary treatment for low-risk patients, across a wide range of clinically relevant threshold probabilities

## Discussion

In this study, we developed and validated the AB-IPI, a prognostic model integrating tumor burden, host fitness, and tumor biology. By incorporating serum albumin and BCL2 expression, the AB-IPI addresses the limitations of the standard IPI in the rituximab era^12^.

The identification of hypoalbuminemia as a strong prognostic factor (HR 2.62) warrants attention. While often considered a nutritional marker, hypoalbuminemia in lymphoma reflects host-tumor interaction involving systemic inflammation. Inflammatory cytokines such as IL-6, secreted by the tumor microenvironment, suppress hepatic albumin synthesis^41^. Furthermore, albumin and IgG ^22^ share the neonatal Fc receptor (FcRn) salvage pathway, which protects them from catabolism. In patients with significant hypoalbuminemia, this recycling mechanism may be impaired, potentially leading to accelerated clearance of rituximab and pharmacokinetic failure, independent of tumor burden. BCL2 overexpression (> 50%) addresses the “Tumor Biology” component, representing an intrinsic barrier to chemotherapy-induced apoptosis^32 30^. Our analysis confirms that this protein-level resistance operates independently of tumor burden.

The approval of Pola-R-CHP has shifted the treatment landscape^16^. However, the POLARIX trial subgroup analysis indicated that the benefit of Pola-R-CHP in patients with an IPI score of 2 was not statistically significant, creating a clinical dilemma.

The AB-IPI offers a refined tool to guide this decision^16^. Our model effectively dissects the intermediate-risk population. Patients with IPI score 2 who are reclassified as AB-IPI High Risk (due to hypoalbuminemia or BCL2 overexpression) represent a population with “compounded resistance” who are unlikely to be cured by R-CHOP and may be the optimal candidates for Pola-R-CHP. Conversely, those reclassified as AB-IPI Low Risk demonstrate excellent outcomes with R-CHOP alone, sparing them the potential toxicity and cost of intensified regimens. The AB-IPI effectively “triages” the intermediate IPI patients, allowing for more precise allocation of novel agents. The AB-IPI is not merely a prognostic calculator but a potential stratification tool for future clinical trials. By identifying ‘true’ high-risk patients within the standard IPI intermediate group, researchers can enrich trials with patients who are most likely to benefit from novel agents like bispecific antibodies or CAR-T cells, thereby maximizing the statistical power of such trials.

### Limitations

This study has several limitations. First, its retrospective nature and single-country cohort (Japan) may limit generalizability, particularly given the higher prevalence of non-GCB subtypes compared to Western populations. Second, while we validated the model using rigorous bootstrapping in accordance with TRIPOD guidelines, the lack of an independent external validation cohort remains a limitation. While our approach provides robust internal validity, confirming the transportability of the AB-IPI in geographically and ethnically diverse populations is a necessary future step. Although external validation is the gold standard, recent methodological guidelines (TRIPOD) emphasize that rigorous internal validation via bootstrapping is superior to split-sample validation in moderate-sized datasets, as it avoids the inefficiency of data discarding and provides stable estimates of model optimism. Our optimism-corrected C-index confirms the robustness of the AB-IPI within this population. Although external validation is widely considered the gold standard, strictly splitting a moderate-sized dataset into development and validation cohorts can reduce statistical power and lead to unstable model estimates. Therefore, we prioritized rigorous internal validity using bootstrapping (1,000 resamples) in accordance with TRIPOD Type 1b guidelines. Recent statistical methodology studies demonstrate that for datasets of this magnitude, bootstrapping provides a more stable and bias-corrected estimate of model performance than split-sample validation, effectively utilizing the full cohort to maximize the reliability of the derived prognostic score, Third, BCL2 expression was assessed by local pathologists without central review; however, the use of a standardized cutoff (>50%) and antibody (Clone 124) mitigates this variability. Direct measurement of serum rituximab levels was not performed, so the link between hypoalbuminemia and pharmacokinetic failure remains inferential in this cohort.

## Conclusion

Despite the lack of statistical significance in discrimination improvement, the AB-IPI demonstrates superior calibration and clinical utility compared to the standard IPI. By integrating markers of host physiology and intrinsic tumor resistance, it offers a practical tool to identify patients who are unlikely to be cured by R-CHOP alone, guiding the optimized use of novel agents.

## Data Availability

This study was conducted in accordance with the Declaration of Helsinki and the Ethical Guidelines for Medical and Health Research Involving Human Subjects in Japan. The protocol was approved by the Ethics Committees of Saitama Medical University Saitama Medical Center (Approval No. 2025-114) and Otemae Hospital. Due to the retrospective nature of the study utilizing existing medical records, the requirement for written informed consent was waived by the Ethics Committees. Instead, an opt-out method was employed, where information regarding the study objectives and exclusion procedures was publicly posted on the institutions' websites and in outpatient clinics, guaranteeing patients the right to refuse participation at any time. Patient records were anonymized prior to analysis. The datasets generated and/or analyzed during the current study are not publicly available due to ethical restrictions mandated by the Ethics Committees of Saitama Medical University and Otemae Hospital, and legal restrictions under the Act on the Protection of Personal Information (Act No. 57 of 2003, amended in 2015) in Japan. The data contain sensitive patient information (e.g., detailed clinical course, admission dates, rare comorbidities) collected under an opt-out consent process, and public sharing could compromise participant privacy. Access to the de-identified minimal dataset is available to qualified researchers upon reasonable request and subject to approval by the Ethics Committee. Requests for data access should be directed to the Ethics Committee Office at

## Author Contributions

N.S. and Y.Tanaka conceived the study, analyzed the data, and wrote the manuscript. K.N. and Y.Takahashi collected clinical data and performed statistical validation. S.M. and M.H. performed the pathological review and BCL2 immunohistochemical scoring. T.T. supervised the study and critically revised the manuscript. All authors reviewed and approved the final manuscript.

## Acknowledgements

We thank the medical staff at Saitama Medical University Saitama Medical Center and Otemae Hospital for their support in data collection.

